# Shift, spark, unfolding: a qualitative life course perspective on the transition surrounding unexpected pregnancy in a Dutch urban setting

**DOI:** 10.64898/2026.03.27.26348213

**Authors:** Merel Sprenger, Matty Crone, Jessica C. Kiefte-de Jong, M. Nienke Slagboom

**Affiliations:** Health Campus The Hague/Department of Public Health and Primary Care, Leiden University Medical Center, The Hague, the Netherlands. Spui 5, 2511 BL The Hague, the Netherlands; Department of Health Promotion, Research Insitutes CAPHRI and NUTRIM, Maastricht University, Maastricht, the Netherlands. Peter Debyeplein 1, 6229 HA Maastricht, the Netherlands.

**Keywords:** Life course, transition, unintended pregnancy, unplanned pregnancy, mothers, fathers

## Abstract

While pregnancy intentions are increasingly recognised as complex and dynamic, unexpected pregnancies are often studied cross-sectionally, and a life course perspective is lacking. This study aimed to explore the salient themes and patterns in 1) the life course trajectories of individuals experiencing an unexpected pregnancy and 2) the transition surrounding an unexpected pregnancy. We conducted semi-structured qualitative interviews with 22 individuals (15 women and 7 men) experiencing unexpected pregnancies. Two interviews were held: during pregnancy and six months after childbirth. The respondents’ life course trajectories were mapped using visual timelines and the transition was explored using Schlossberg’s transition theory. Interviews were thematically analysed in an iterative process: applying open coding to three interviews followed by thematic coding and comparison of themes within and across life course trajectories. Life course trajectories varied considerably, distinguished by the absence or presence of critical life events, with patterns characterised by clustering life events within the domains of family, mental health or frequent residential mobility. The transition process of unexpected pregnancy was characterised by three patterns of adjustment – *shift* (instant adjustment), *spark* (triggered adjustment) and *unfolding* (ambiguous adjustment) – in which respectively, all life course trajectories, trajectories with clustering of life events and mainly stable trajectories were present. This study contributes to the literature through the *unfolding* pattern, showing that adjustment to unexpected pregnancy may be an ambiguous process that is not finished when the baby has arrived, especially if individuals have strong aspirations in light of a relatively stable life course.

**Highlights:**

- Unique in unintended pregnancy research: life course, during pregnancy and postpartum.
- Life course shows (in)stability regarding family, mental health and residential mobility.
- Three patterns in adjusting to unexpected pregnancy: instant, triggered and ambiguous.
- Adjustment and ambiguous feelings may extend beyond pregnancy.
- Ambiguous adjustment mainly included people with stable life course trajectories.

## Introduction

While pregnancy intention is increasingly recognised as complex and dynamic (Auerbach et al., 2023; Helfferich et al., 2021), most qualitative research on unexpected pregnancy^1^ continues to focus on a single point in time. This limits the understanding of how unexpected pregnancy is embedded within people’s lives and how it is experienced over time. Only two qualitative studies have explicitly conceptualised unexpected pregnancy as a *transition* (Aldrighi et al., 2024; Gbogbo, 2020), broadly defined as “any event or nonevent that results in changed relationships, routines, assumptions, and roles” (Anderson, 2012, p. 39). Aldrighi et al. (2024) studied high-risk unplanned pregnancies at an advanced maternal age in Brazil and found that emotional and psychosocial complexity, particularly relating to stigma and challenges relating to older age, interfere with this transition. Gbogbo (2020) studied adolescent women who had experienced or were experiencing unplanned pregnancies in Ghana, highlighting challenges related to their age and financial insecurity, alongside the presence of some social support.

Following these two studies, the present study also applies Schlossberg’s Transition Theory (Aldrighi et al., 2024; Anderson et al., 2012; Gbogbo, 2020; Schlossberg, 1981). In this theory, Schlossberg considers an individual’s potential resources – their assets and liabilities – for coping with the transition, providing a framework for studying transition processes such as pregnancy by mapping four major sets of factors referred to as the four S’s: situation, self, support, and strategies. Factors relating to the situation give information on what is happening and in which context, including previous experiences, it is occurring. Self includes personal characteristics such as socioeconomic status, age, and gender, but also considers one’s psychological resources including values and outlook on life. Support identifies what types of support – emotional support, affirmation, or practical help – people may access and from whom they may receive it, such as family, friends, coworkers, neighbours, or others. Finally, strategies addresses how individuals may cope instrumentally, i.e., by changing or taking control of the situation, or emotionally, i.e., by managing stress (Anderson et al., 2012; Schlossberg, 1981). Although Gbogbo (2020) and Aldrighi et al. (2024) brought about relevant new insights, particularly relating to unexpected pregnancy as a transition, they focused on specific populations – either younger or older individuals – and were conducted in low- and middle-income countries (Aldrighi et al., 2024; Gbogbo, 2020).

Some qualitative studies in high-income countries have suggested that the experience and impact of unexpected pregnancies are shaped by individuals’ circumstances and life stages at the time of pregnancy (Helfferich et al., 2014; Kavanaugh et al., 2017). In the United States, Kavanaugh et al. (2017) retrospectively examined how unintended childbearing affected individuals’ education, career, finances, relationships and health, showing that impacts varied according to workplace flexibility, government assistance, interpersonal support, pregnancy or birth-related health and substance use. Similarly, a study in Germany demonstrated that the circumstances associated with unintended pregnancy differed by age and life stage, including educational enrolment, one’s career, relationship status and whether individuals already had children (Helfferich et al., 2014). Together, these studies suggest that unexpected pregnancies cannot be understood in isolation from people’s current life situations. However, their primary focus remains on where in the life course individuals are, i.e. at which age or life stage, at the moment of pregnancy rather than on the broader trajectories preceding the pregnancy.

A life course perspective may offer a broader understanding of unexpected pregnancy by situating it within diverse life trajectories. This perspective is particularly valuable given evidence that transitions to parenthood are shaped not only by current circumstances but also by earlier life events, both adverse and advantageous (Mathijssen et al., 2024). Adversity in early life has been shown to contribute to increased health-risk behaviours, including sexual risk behaviour, as well as poorer physical, mental and social health outcomes in adulthood, although individuals may also develop resilience despite having experienced adversity (Hughes et al., 2017; Kim & Royle, 2025; Leung et al., 2022; Petruccelli et al., 2019). A recent meta-analysis further demonstrated an association between adverse childhood experiences and unintended pregnancy, with particularly strong effects for experiences related to family dysfunction and sexual abuse (Li et al., 2025). Despite this, to the best of our knowledge, no prior qualitative study has examined unexpected pregnancies from a life course perspective.

To conclude, knowledge remains limited regarding life course trajectories prior to unexpected pregnancies and regarding transition experiences of unexpected pregnancies, especially in the general population of high-income countries. Moreover, while the importance of partners and men is increasingly recognised (Van der Sluis et al., 2015; Wittig & Rodriguez, 2019), research on unintended pregnancies seldom includes their experiences (Chan, 2021; Combs et al., 2021; Lindberg & Kost, 2014; Sayler et al., 2021). In light of these gaps in the literature, this study aims to explore the salient themes and patterns in 1) the life course trajectories of women and men experiencing an unexpected pregnancy and 2) the transition surrounding unexpected pregnancy.

## Methods

### Study design and context

This qualitative life course interview study was embedded in a larger mixed methods study on unexpected pregnancy called the risk and resilience of unintended pregnancy (RISE UP) study. In addition to the qualitative interview study presented in this paper, the RISE UP study included a quantitative survey component (Sprenger, Crone, et al., 2025). The interview study was exempted from review by the [name of review committee, reference number]. The study is set in the Hague, a city in the Netherlands with around 566,000 inhabitants in 2024 and around 5,500 births in 2023 (Gemeente Den Haag & DSO /SEPO - Onderzoek, 2024). In the Netherlands, pregnancy care is primarily organised by primary care midwives or, if specialised care is needed, by gynaecologists or clinical midwives in the hospital (Rijksinstituut voor Volksgezondheid en Milieu, 2025).

### Inclusion criteria and recruitment

Respondents were approached after completing the RISE UP study survey. In this survey, pregnant people and their (ex-)partners completed the Dutch Adaptation of the London Measure of Unplanned Pregnancy (DA-LMUP), an instrument that measures pregnancy intention on multiple dimensions (Barrett et al., 2020; Barrett et al., 2004; Sprenger, Beumer, et al., 2025). Inclusion criteria to enter the interview study were to a) live in The Hague and have a non-intended pregnancy as indicated by a DA-LMUP score of ≤9 on the RISE UP study survey, or be a (current or former) partner of such a person (Hall et al., 2017); b) be aged 16 years or older; and c) speak Dutch, Turkish, Arabic, Mandarin, Polish, Bulgarian or English. First, the researcher invited potential respondents by sending emails with information about the interviews. Their partners were also invited to participate in an interview and to complete the RISE UP survey if they had not already done so. If they had not denied participating, the researcher would call them one week later, answering any questions, providing further information, if needed, and scheduling the first interview if they wanted to participate. Respondents were sent the informed consent form prior to the interview and signed it before the start of the interview.

### Data collection

The study included two rounds of semi-structured life course interviews: one interview during pregnancy and the other 6 months after childbirth. The topic guide for the interviews can be found in the RISE UP study project on the Open Science Framework: https://doi.org/10.17605/OSF.IO/63HBE. All interviews were performed in Dutch or English and took place at the respondents’ places of choice, often at their homes. The interviews were recorded and transcribed verbatim. The interview data are confidential. Interviews were conducted by the first author, who reflected on the process in a journal. Data collection ended when saturation was reached (Frambach et al., 2013). The first interview, individually conducted with 22 respondents (15 pregnant people and 7 of their partners), focused on the life course trajectory and experience of unexpected pregnancy during pregnancy.

Before starting the interview, sociodemographic characteristics, such as age, neighbourhood in the Hague, relationship status, education, employment and children, were collected (Table 1). In the interview section regarding the life course, we explored the life course trajectories of the respondents from birth to the present. The topic guide included questions on childhood and family, education and employment experiences, experiences with care and their experiences with relationships and reproductive life events. For example, we asked, “Can you tell me how it was growing up when you were young?” and “How and when did you learn about relationships, for instance, at home?” Meanwhile, the interviewer wrote down all life events on a timeline. Timelines can help uncover tacit knowledge; they improve rapport, decrease the hierarchy between interviewer and respondent, can be used to triangulate and are useful when talking about sensitive subjects (Adriansen, 2012). When the timeline arrived at the pregnancy, the interviewer asked the respondent whether the timeline matched their story and whether there were any important life events that were missing, needed adjustment, or required addition in the timeline.

**Table 1.** Characteristics of respondents in the first interview, presented as mean [range] or n (%)

| **Characteristic** | **All respondents (n=22)** | **Pregnant women (n=15)** | **Partners (n=7)** |
| --- | --- | --- | --- |
| Age | 30.6 [20-40] | 30.7 [20-40] | 30.6 [20-38] |
| Education level |  |  |  |
| Secondary (vocational) | 5 (22.7) | 3 (20.0) | 2 (28.6) |
| Tertiary | 17 (77.3) | 12 (80.0) | 5 (71.4) |
| Religious |  |  |  |
| No | 15 (68.2) | 11 (73.3) | 4 (57.1) |
| Yes | 7 (31.8) | 4 (26.7) | 3 (42.9) |
| Migration background |  |  |  |
| No | 11 (50.0) | 9 (60.0) | 2 (28.6) |
| Yes | 11 (50.0) | 6 (40.0) | 5 (71.4) |
| Relationship status |  |  |  |
| Single | 2 (9.1) | 2 (13.3) | 0 |
| Cohabiting or married | 20 (90.9) | 13 (86.7) | 7 (100.0) |
| Children |  |  |  |
| 0 | 11 (50.0) | 7 (46.7) | 4 (57.1) |
| 1+ | 11 (50.0) | 8 (53.3) | 3 (42.9) |
| Gestational age (in weeks) | 26.9 [10-36]<sup>a</sup> | 24.9 [10-36] | 31.7 [24-36]<sup>a</sup> |
| Pregnancy intention score<sup>b</sup> | 6.3 [1-11]<sup>b, c</sup> | 6.1 [1-9] | 6.8 [2-11]<sup>c</sup> |
| Adverse life events, median (IQR)<sup>b</sup> | 3 (1.75-6) | 3 (1.5-6) | 3 (2-4) |
| 0 | 0 | 0 | 0 |
| 1 to 3 | 12 (54.5) | 9 (60.0) | 3 (42.9) |
| 4 or more | 8 | 6 (40.0) | 2 (28.6) |
| Missing | 2 | 0 | 2 (28.6) |

<sup>a</sup> One interview with a partner was conducted three weeks after birth; gestational age included for six partners.
<sup>b</sup> As measured in the RISE UP study survey.
<sup>c</sup> Two partners did not complete the RISE UP study survey; pregnancy intention scores included for five partners.

In the interview section regarding pregnancy, we explored how the respondents experienced the transition surrounding the pregnancy. This part of the interview guide was based on Schlossberg’s Transition Theory stress (Anderson et al., 2012; Schlossberg, 1981). Questions included, for example, “How did you feel when you found out about the pregnancy?”, “How did your family respond to the pregnancy?” and “How have any life events we discussed earlier impacted the pregnancy?”

In the second interview round, at 6 months after childbirth, 13 of the 15 women and 8 partners (all and one who did not participate in the first interview) participated. Couples were interviewed together, resulting in 14 interviews, of which 7 were with couples. The primary objective of this interview differed from the objectives of the current manuscript, as it focused on support and care needs and experiences. For this manuscript, the relevant section of the second interview was that in which respondents were asked to reflect on the unexpected nature of the pregnancy.

### Analysis

The interview data were thematically analysed to identify salient themes and patterns, following Braun and Clarke’s (2006) approach. To explore salient themes and patterns in the life course trajectories of individuals experiencing an unexpected pregnancy (objective 1), the analysis followed three steps. First, the first three life course interviews were coded inductively and independently by three researchers, after which codes were compared and discussed to enhance analytic depth. Second, visual life course trajectories were constructed for all respondents across three life stages: childhood (_≤_12 years), adolescence (12–18 years) and adulthood (_≥_18 years). Life events were categorised into life domains: family, health, violence, education, bullying, mobility, reproduction, and interactions with legal or governmental institutions. Third, life trajectories and themes across respondents were systematically analysed.

To address the second objective, exploring patterns in the transition surrounding an unexpected pregnancy, transcripts from the second interview, conducted approximately six months after the pregnancy, were analysed, building on Schlossberg’s Transition Theory (Anderson et al., 2012; Schlossberg, 1981). Coding focused on how the respondents described navigating the transition with regards to their coping resources: those relating to the situation (e.g. timing, role change and concurrent stress), self (e.g. personal characteristics and psychological resources), support (e.g. from partner, family, friends and work) and strategies (e.g. to manage stress or to modify or control the situation). In our analysis of the transition, it stood out how the respondents primarily referred to a certain adjustment taking place, guiding our analysis to focus on different patterns of adjustment. Finally, we compared these adjustment patterns across types of life course trajectories. For validation, findings were discussed in a meeting with health care providers and policy advisors in The Hague who confirmed the identified patterns resonated with their observations and experiences as professionals.

## Results

Table 1 shows the characteristics of the 22 respondents, of which 15 were pregnant and 7 were their current partners. They were, on average, 31 years old (range 20–40 years). Most of them were not religious (68.2%) and had a tertiary education (77.3%). The gestational age at the first interview ranged from 10 to 36 weeks, with a mean of 27 weeks.

### Life course trajectories

The first objective of this study was to explore salient patterns across the life course trajectories of individuals who experienced unexpected pregnancies. Although life courses varied considerably, two key distinctions emerged. The first concerned the volume of life events, that is, the degree of instability to which the respondents were exposed prior to their pregnancies, especially before the age of 18, resulting in two groups: those with a relatively stable life course trajectory (7 respondents) and those who had experienced more instability (15 respondents). Stable life course trajectories showed more similarity than life trajectories with more instability. In the stable group, all respondents recounted stability across various domains, including at home and at school. For example, one respondent (F14) described that her parents “always knew what was going on, so it felt very adventurous, but it was very safe”, and F1 described her educational trajectory as “passing with flying colours, easily” (F1). Within life course trajectories with more instability (i.e. more adverse life events or transition events), a second distinction could be made regarding the nature of the life events, revealing three distinct patterns, primarily characterised by life events in the domains of mental health (n = 5), family (n = 5) and residential mobility (n = 5). These three patterns of instability are elaborated upon below.

### Mental health

Within the first pattern of instability, the respondents experienced clustering of life events related to mental health, both of their family members (parents or siblings) and themselves. Parental mental health problems included depression, borderline personality disorder, addiction, and attention deficit hyperactivity disorder (ADHD). Sibling’s difficulties included autism or anger problems. Growing up in a home with family members struggling with their mental health disrupted family life and reduced attention for respondents as they were growing up. As adults, some respondents struggled with different conditions such as epilepsy, sleep problems, depression, and anxiety. Respondent F5, for example, described the instability she grew up in by saying, “My mother has a lot of mental issues, so there was always a lot of attention for that. […] She has a very typical borderline personality, I would say, very fickle and not knowing where you stand.”

As a young adult, F5 suffered from anxiety, and later, she was diagnosed with ADHD. Similarly, M6 recounted how his brother’s behaviour affected him and his parents throughout his childhood, as he would get “quite violent and hit people and bite people and try and slam their fingers in doors”. M6 himself suffered from social anxiety after changing schools at a young age and had to deal with chronic back pain since his late teenage years.

### Family

Adverse events in the family environment marked the second pattern of instability, in which the respondents described growing up amid parental tensions and divorce. The respondents remembered conflicts and ensuing instability, recollecting the impact of the tensions both before and after the divorce. For example, F4 described her parents’ unstable relationship and looming divorce impacting her wellbeing as a child; she said, “I really struggled with it. I was often crying at school and at friends’ houses, because I kept thinking my parents were going to separate.”

When parents eventually divorced, it was a relief for some, but the tensions and instability around the divorce could also continue to impact them long afterwards. This is illustrated in F7’s life course trajectory, in which she spoke about her parents fighting throughout her childhood and the limited contact with her father after the divorce during her teenage years: “I had an anxiety disorder, partly because of that. I was very shy and insecure for a long time, and often quite withdrawn. That lasted until I was in my mid to late twenties.”

Reduced contact or broken contact with a parent or sibling, sometimes temporarily and indefinitely at other times, was another source of instability, which was experienced by three respondents. F15, for example, when her parents’ separation was definite after they had been on and off for a while, recalled, “When I was fourteen, I said, ‘I am so done with it all and I don’t need to see him anymore, because he is not a normal father and it’s not doing me any good.’”

### Residential mobility

Frequently moving from one place to the other – often between countries, either due to parents’ jobs or migration in search of a better life – introduced considerable instability into the early life course. Parental employment, for example, working for international organisations or the military, led to families having to move along to the parents’ new workplace location. M2, for example, who moved frequently because his father worked in the military service, reflected on the moves – he attended over ten different schools across South and West Asia – as introducing instability before getting used to the new situation, saying, “There was a lot of shifting in the beginning then towards the end, it became stable.” He recounted feeling sad about moving – “you make friends, and you leave” – he still does not get incredibly attached to things or people.

In another example, F9 (age range 20-24) grew up in a family searching for “better opportunities, better social insurance, better health insurance and […] better Islamic groups”. She recollected residential mobility among her country of birth in North Africa, the United Kingdom, and the Netherlands, before settling there, shifting between different cultures, not being at the same school for more than two years and supporting her parents with navigating Dutch institutions. The pattern of residential mobility was continued by all respondents in their adult lives.

### Transition of unexpected pregnancy

After exploring life course trajectories, the section below attends to patterns in the transition surrounding unexpected pregnancies. An important note is that although all the respondents expressed a desire for (more) children in the future, just not now: none of them were actively trying to conceive. Regarding the transition, all the respondents described a certain adjustment taking place. For some, this adjustment was an immediate moment, the instant of discovering they found out about the pregnancy (shift); for some, it was triggered at a later moment (spark), and for others, it was not a moment, but a, perhaps still continuing, process over time (unfolding). This is why we do not speak of acceptance associated with an endpoint but with adjustment, which may represent both a moment and a continuing process (Ferguson, 2022; Purdy, 2024). These patterns of adjustment, presented in Table 2, are discussed below alongside key moments in time and illustrative quotes.

**Table 2.**
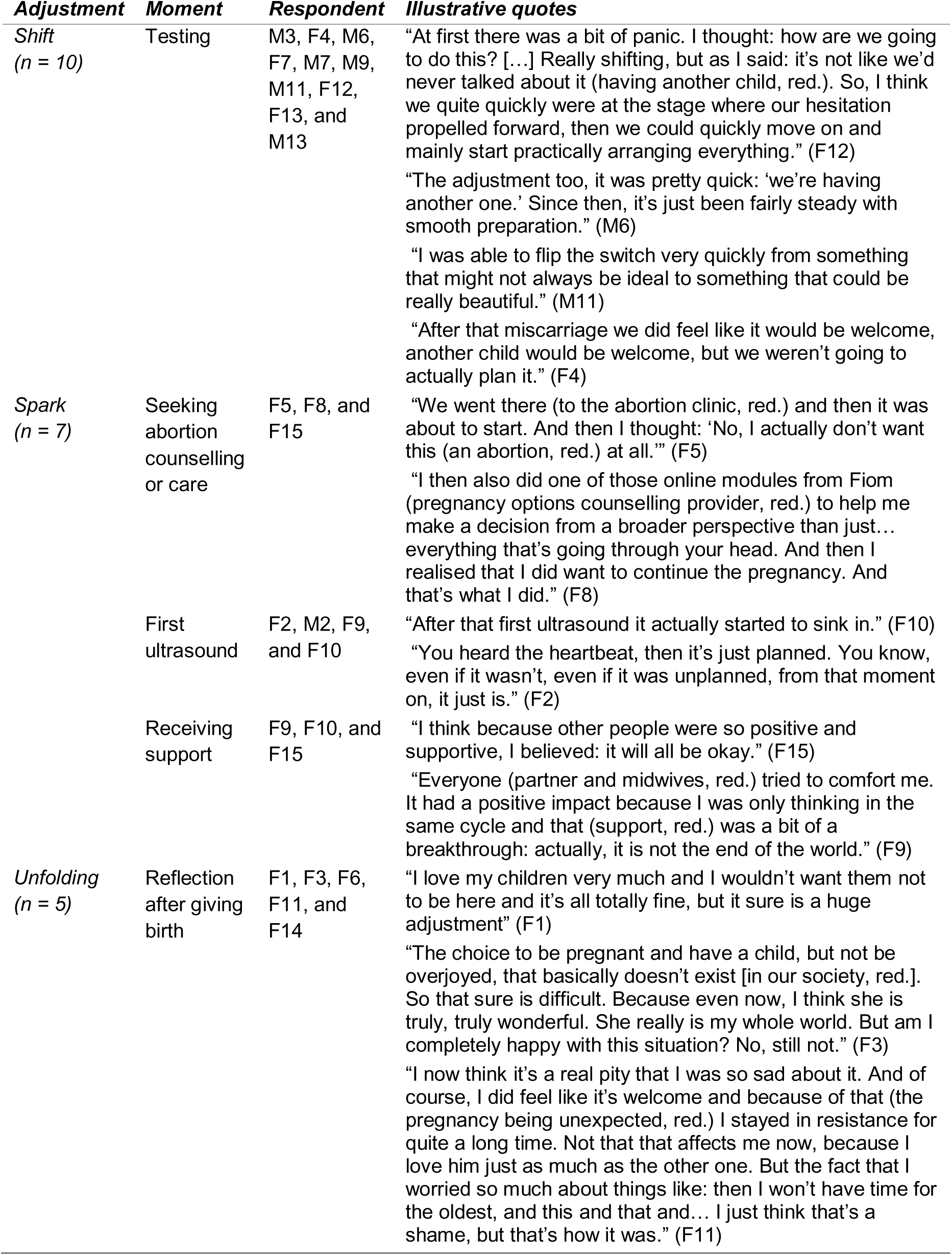
Moments of adjustment.

For most of the respondents, the option of abortion briefly crossed their minds or was discussed with their partners. For example, Z10’s partner told her, “If you do not want to keep it, I will support you.” However, although she felt unsure about the pregnancy and raising a child, abortion was not an option for her. Like Z10, the decision to continue the pregnancy was sometimes (partially) grounded in the ‘unchoosing’ of abortion (Kimport, 2022), where abortion was not an option due to preconceptions relating to, for example, religion or an earlier embodied abortion experience. This ‘unchoosing’ of abortion was present across all life course trajectories and adjustment patterns, although for women, it was more present for the spark and unfolding adjustment patterns.

### Shift: Instant adjustment

As illustrated in Table 2, a recurring feature in this pattern of adjustment is how the respondents described their instant responses upon discovering the pregnancy. After testing positive for pregnancy, they recollected “flipping the switch” (M11 / age range 35-40; M9 / age range 20-24) or “shifting” (F12 / age range 30-34). This went hand in hand with positive emotions, such as excitement and happiness; for some, it was accompanied by brief feelings of shock or panic. This adjustment pattern occurred across all life course trajectories. Notably, all but one interviewed partner (all men) adjusted in this way. For example, M3 (age range 25-29) was “instantly happy” and explained that “there was a shift, we are now entering the adult life”.

In another example, M7 (age range 25-29) felt “excited about the fact that we’re going to have a kid – it’s a little early, but we were planning on trying in nine months anyway”. Similar to other respondents who recollected instant adjustments, M7 had a stable livelihood. He and his wife were open to having a baby. The addition of a child to their lives did not seem to affect their life aspirations; it simply moved up the timeline. When reflecting on the unexpected nature of the pregnancy during the second interview, the respondents with this adjustment pattern most often said that the unexpected nature of the pregnancy was no longer important. M7 explained, “There could have been a better time, but I also didn’t really think it matters.” Two respondents who already had a child formed an exception to this, saying that, although they did not regret having the child and were happy with this addition to their family, “objectively, we would probably be happier at this stage if we only had one child compared to two children” (M6 / age range 35-39).

### Spark: Triggered adjustment

Respondents from this adjustment pattern were in shock when they discovered the pregnancy and experienced excitement and fear. For example, F5 said, “I was very happy, but at the same time also felt scared. I thought, shit, how do I do this?” (F5 / age range 30-34). As illustrated in Table 2, for this adjustment pattern, there were three main pivotal moments sparking adjustment, described by respondents with life course trajectories with some degree of instability (mental health, family, and residential mobility). Most respondents within this pattern had been in a relationship for a short time or were experiencing relational instability when they found out about the pregnancy. They all had a desire to have children but did not have a clear moment in mind.

One pivotal moment could be when the respondents considered – but did not choose – abortion, as for F5: “We went there (to the abortion clinic, red.) and then it was about to start. And then I thought: ‘No, I actually don’t want this (an abortion, red.) at all’” (F5). Through the process of considering abortion, these respondents realised that they actually did want to have babies. Another pivotal moment could be the first ultrasound, confirming that the pregnancy was actually there, for a woman aged 25-29 resulting in reframing: “You heard the heartbeat, then it’s just planned. You know, even if it wasn’t, even if it was unplanned, from that moment on, it just is” (F2). Finally, upon receiving family and peer support after breaking the news of pregnancy and its unexpected nature, a pregnant woman reflected that it “really does feel like you’re not on your own” (F10, age range 25-27). Support served as comfort; it helped the respondents believe that, on the one hand, they could be parents and, on the other hand, people around them would support them. When reflecting on the unexpected nature of the pregnancy six months afterwards, the respondents were generally happy or found it irrelevant: “This is our story, at a certain moment it is not even that interesting anymore whether she was planned or not, she is just here.” (F5)

### Unfolding: Ambiguous adjustment

Respondents in this pattern, all women, primarily with a stable life course trajectory, expressed continued difficulty coping with the pregnancy. Discovering the pregnancy was often experienced as surreal, evoking feelings of anger, sadness, panic, denial, or shame. For example, a female respondent remembers thinking “now, my life is over.” (F3, age range 25-29) Another female respondent explained her feelings as follows: “A bit angry, a bit fed up, a bit sad, a bit angry, a bit of everything all at once. Because the opposite would be being completely overjoyed. So, that makes it a bit difficult.” (F1, age range 30-34) These initial feelings resonated strongly during the first round of interviews conducted at the gestational age of 15–32 weeks. The responses to the unexpected pregnancy were often related to the incompatibleness of having a baby with current aspirations in life. Each of the respondents wanted to have more time in their current situation before adding a(nother) member to the family and focus on themselves (e.g. their career or wellbeing) or spend time with the child they already had. For example, F3 was pursuing an international career and was just about to move abroad for work. Because of the pregnancy, she could not do this:

> *I think I really had to pull myself together a bit, because I was very focused on my career first, on doing what I wanted. And when that suddenly goes completely in the opposite direction, that’s quite a disappointment. And besides that, I had a lot of energy. I exercised a lot, really a lot, and within two months that completely disappeared. So also, you know, that sense of having no control, that things can change so drastically in such a short time. (F3)*

During the second interview six months after giving birth, the respondents reflected on the unexpected pregnancy and now having a(nother) child, describing that their feelings had partially changed. As shown in Table 2, the respondents expressed ambiguity in their reflections after birth. While they were happy with their children, they also continued to feel challenged. One respondent, for example, said, “She really is my whole world. But am I completely happy with this situation? No, still not.” (F3) Others were happy but thought it was a pity that they had had such difficulty being at peace with the pregnancy, especially knowing what they knew now:

> *Yeah, an unplanned pregnancy, it’s… And I think you can have that with a planned one, too, but with a spontaneous one it’s really… It hits you in the face. Immediately, there’s a timer hanging over your head: nine months, there you go. […] If I had known back then that it would be her who would arrive, I probably would have been much more at ease. Oh, the baby sleeps, she poops, she eats, she smiles all the time. She thinks everyone is absolutely lovely. If someone had told me that I would get a baby like that, I would have said: alright then. (F14, age range 30-34)*

It stands out that this type of adjustment shown by these women contrasted with how their partners adjusted to the pregnancy – they shifted immediately. For example, F3’s partner adjusted immediately to the pregnancy, whereas she experienced ambivalence, so when “[she] really thought like [she] actually totally doesn’t want this […] it was difficult to complain about this to him” (F3). Nevertheless, her partner’s more positive attitude towards the pregnancy was helpful, too: “If he would not have wanted it either, then [they] would have been a bit down and out together, but that (him being positive red.) was actually really nice” (F3).

## Discussion

In this study, we explored the salient patterns in the life course trajectories of individuals with unexpected pregnancies, as well as the transition surrounding unexpected pregnancies. In the respondents’ life course trajectories, we found patterns based on the volume of adverse life events they had experienced growing up and the type of adversity relating to family, mental health, or residential mobility. With regards to the transitional phase around an unexpected pregnancy, the respondents discussed their path of adjustment. We identified three patterns of adjustment: instant, triggered and ambiguous. When asked for a reflection on their current pregnancies in light of their life course trajectories, the respondents reflected on their prospective parenthood instead. Nevertheless, we found a pattern along the identified life course trajectories and adjustment patterns. All life course trajectories were present in the *shift* pattern; the *spark* pattern included respondents who had experienced instability (regarding family, mental health, or residential mobility), and the *unfolding* pattern applied mainly to those with a stable life course trajectory.

The *shift* and *spark* patterns largely align with two types identified in a typology of emerging acceptance in unintended pregnancies described in a German mixed methods study, respectively: ‘acceptance before conceiving’ relating to a generalised readiness for having a baby and somewhat consciously taking a chance by not (always) using contraception and ‘acceptance due to a turning point’ relating to a trigger such as seeing an ultrasound image (Helfferich et al., 2021). Regarding the *spark* pattern, the pivoting moment of support was also found to facilitate the transition around unintended pregnancy in Brazil (Aldrighi et al., 2024). In our sample, some respondents’ preconceptions of abortion not being an option align with work in the United States, where, for some, abortion was ‘unchoosable’ as a result of cultural beliefs, experience and embodiment or structural barriers (absent here, potentially reflecting Dutch accessibility to abortion). Others considered abortion but chose pregnancy because they wanted a baby (Kimport, 2022).

Although the concept of ambivalent feelings around a pregnancy has long been discussed (Auerbach et al., 2023; Cutler et al., 2018; Klerman, 2000; Schwarz et al., 2007), this study is among the first to examine adjustments to unexpected pregnancy both during pregnancy and six months postpartum. Whereas the typology by Helfferich et al. (2021) described one type of acceptance that eventually emerged during pregnancy after feelings of ambivalence, our *unfolding* pattern suggests that this may not always conclude within pregnancy or early parenthood, often tied to conflicting aspirations, echoing a recent Dutch report on circumstances and care experiences around unexpected pregnancy (Van Ditzhuijzen et al., 2025). Similarly, although in a different sample, a mixed methods study in the United States found prevailing mixed emotions among those denied abortion (i.e., carrying an unwanted pregnancy to term), with negative emotions stabilising after giving birth and positive feelings increasing over time – a gradual process, facilitated by instrumental and emotional support (Rocca et al., 2021).

Two additional striking observations emerged regarding the adjustment patterns. First, the presence of stable life course trajectories within the *unfolding* pattern – often accompanied by conflicting aspirations, as has been identified in earlier studies (Aldrighi et al., 2024; Brauer et al., 2019; Spierling & Shreffler, 2018; Yong et al., 2023) – raises the question of why those with relatively stable lives experienced more ambiguous adjustment. One potential explanation might be that some exposure to adversity could foster resilience in coping with new, unanticipated life events, whereas too little or too much does not (Carstensen et al., 2020; Seery, 2011). When someone has experienced and overcome adversity, they may be more inclined to think that they can also overcome a new unanticipated life event, such as an unexpected pregnancy. Nevertheless, both the volume and timing of life events are relevant and intersect with broader social conditions, such as socioeconomic status, gender and race or ethnicity (Comolli et al., 2024; Hall et al., 2019). For example, while women generally experience more critical life events, concentrated events have a greater impact on men; when events are more broadly distributed across the life trajectory, the effects are similar across genders (Comolli et al., 2024). Another potential explanation might be that a more stable life course trajectory fosters a strong internal sense of control – believing that they are responsible for all failures and successes in their life and consequently expecting any challenges to be taken on and resolved (Elder, 1998). An unexpected pregnancy may disrupt this belief, as it likely triggers other transitions rather than fitting into one’s existing life plans. A previous study found that unanticipated events might be more challenging for women with a stronger internalisation of control compared to men (Stillman & Velamuri, 2016).

The second observation concerns gendered patterns of adjustment. Notably, the *unfolding* pattern was observed exclusively among women, whereas nearly all men seemed to adjust immediately. Although not all male partners participated in the study, women’s accounts suggest that this instant adjustment pattern also characterised most non-participating men. This raises the question of why men seem to adjust rapidly, while women display more varied trajectories and are particularly represented in the *unfolding* pattern. One explanation might be that pregnancy – particularly when unexpected – potentially affects those carrying it more profoundly regarding health, wellbeing, economic stability and aspirations (McAuliffe, 2023; Sonfield et al., 2013). Another explanation relates to gendered norms around expressing ambivalence regarding pregnancy and parenthood. In some contexts, as suggested by F3, it may not be societally accepted for women to express these feelings while they are held to heightened expectations of being a ‘good mother’, whereas men may be granted more leniency – although they too are increasingly expected to fulfil an active parenting role (Faircloth, 2023; Kukura, 2025). Supporting this interpretation, a study of young Dutch fathers found that unexpected pregnancy challenged men’s sense of stability and often elicited a pragmatic, resilient stance that may inhibit emotional processing and help-seeking (Van De Beek et al., 2025). The apparent instant shift among men in our study may reflect a similar adaptive response.

### Strengths and limitations

Applying a life course perspective and two interview rounds – during pregnancy and after birth – this study captured life course trajectories before and through unexpected pregnancy, offering a unique view of adjustment as it unfolds. Schlossberg’s Transition Theory provided a solid theoretical foundation for studying transition, while a more open, exploratory approach might have revealed aspects of the transition surrounding unexpected pregnancy that the theory does not currently capture. Including both parents added insight into gendered differences in adjustment. Several limitations should be acknowledged. First, all partners who agreed to participate were in a relationship with the pregnant women at the time of the first interview, limiting the perspectives of men who did not participate, two of which were no longer in a relationship, likely reflecting the experiences of more involved men. Second, due to sampling via the RISE UP study survey and the lack of inclusion of people who had had an abortion, our findings apply only to the experience of pregnancies carried to term. Finally, the sample included predominantly individuals around the age of 30 with a higher socioeconomic position, both with and without a migration background. While this is a strength, given that these groups are often underrepresented in research on this topic, the findings should be interpreted with caution when considering younger individuals and individuals with lower socioeconomic positions.

### Implications

These findings show that the transition to parenthood in the case of an unexpected pregnancy can be a diverse experience for different people. A meaningful approach in care would be to explore how the pregnancy fits into someone’s life course trajectory, current circumstances and life aspirations while recognising that adjustment might, for some, be an ambiguous process that may continue after birth rather than a single moment of acceptance and that within couples, different adjustment patterns may prevail. Further, especially in these cases, it is recommended to discuss adjustment to unexpected pregnancy not merely at the beginning of the pregnancy but to revisit this topic throughout the pregnancy and in the postpartum period – as a care provider, relative or friend. Future research should further explore what types of support people with different adjustment patterns – particularly those with an unfolding pattern – would like to access, and how the extent, type, and timing of adversity in the life course shapes coping with subsequent unexpected or adverse events.

### Conclusion

This study explored the salient themes and patterns in the life course trajectories of individuals experiencing an unexpected pregnancy and in the transition surrounding it. Regarding life course trajectories, patterns emerged based on both the volume of adverse life events experienced growing up and the type of adversity – relating to family, mental health, or residential mobility. Regarding the transition surrounding unexpected pregnancy, three patterns of adjustment were identified – instant, triggered, and ambiguous – showing that for some, adjustment is best understood as a dynamic, ongoing process rather than a single moment of acceptance. Notably, although findings should be interpreted with caution given the small subgroup, those with relatively stable life course trajectories were more likely to experience ambiguous adjustment, suggesting that not only too much, but also too little prior adversity may hinder adaptive coping with unanticipated life events. A further striking finding was that adjustment patterns diverged along gendered lines within couples, with men more frequently adjusting instantly while women displayed more varied trajectories, potentially reflecting the unequal physical, emotional, and social implications of pregnancy. Together, these findings highlight the need for care that moves beyond the question of whether a pregnancy was planned towards a broader understanding of how it fits within people’s life course trajectories and aspirations. Further, they point to the importance of attending to adjustment throughout pregnancy and into early parenthood, including the potentially diverging experiences of both parents within a couple.

## Data Availability

The interview data are confidential.

https://doi.org/10.17605/OSF.IO/63HBE

## Acknowledgements

First, we would like to express our utmost gratitude to the respondents who were willing to share their life stories with us. Further, we are grateful to colleagues at Health Campus The Hague/Department of Public Health and Primary Care (Leiden University Medical Center), the Centre for Biomedicine, Self and Society (University of Edinburgh) and the Advancing New Standards in Reproductive Health Institute (University of California San Francisco) for their thoughtful questions and discussions as we presented our work in progress. This work was supported by ZonMw [grant numbers 554002006; 554001004].

## Declaration of generative AI and AI-assisted technologies in the writing process

During the preparation of this work, the authors used ChatGPT and Claude in order to improve flow. After using this tool, the authors reviewed and edited the content as needed and take full responsibility for the content of the published article.

## Footnotes

1 In this paper, we use the term *unexpected pregnancy*, which encompasses the experiences of all respondents in our study, unlike terms such as unintended or unplanned, which apply only to some. When referring to previous studies, we use the term employed by the authors.

